# Significant impacts of the COVID-19 pandemic on race/ethnic differences in USA mortality

**DOI:** 10.1101/2022.04.04.22273385

**Authors:** José Manuel Aburto, Andrea M. Tilstra, Ginevra Floridi, Jennifer B. Dowd

## Abstract

The COVID-19 pandemic triggered declines in life expectancy at birth around the world. The United States of America (USA) was hit particularly hard among high income countries. Early data from the USA showed that these losses varied greatly by race/ethnicity in 2020, with Hispanic and Black Americans suffering much larger losses in life expectancy compared to white people. We add to this research by examining trends in lifespan inequality, average years of life lost, and the contribution of specific causes of death and ages to race/ethnic life expectancy disparities in the USA from 2010 to 2020. We find that life expectancy in 2020 fell more for Hispanic and Black males (4.5 years and 3.6 years, respectively) compared to white males (1.5 years). These drops nearly eliminated the previous life expectancy advantage for the Hispanic compared to white population, while dramatically increasing the already large gap in life expectancy between Black and white people. While the drops in life expectancy for the Hispanic population were largely attributable to official COVID-19 deaths, Black Americans additionally saw increases in cardiovascular disease and “deaths of despair” over this period. In 2020, lifespan inequality increased slightly for Hispanic and white populations, but decreased for Black people, reflecting the younger age pattern of COVID-19 deaths for Hispanic people. Overall, the mortality burden of the COVID-19 pandemic hit race/ethnic minorities particularly hard in the USA, underscoring the importance of the social determinants of health during a public health crisis.

**Significance statement:** Public interest in social and health inequalities is increasing. We examine the impact of COVID-19 on mortality in the USA across racial/ethnic groups and present four key findings. First, all groups suffered sizable life-expectancy losses and increases in years of life lost. Mortality from cardiovascular diseases, “deaths of despair”, and COVID-19 explained most of these losses. Second, working-age mortality accounted for substantial life-expectancy losses, especially among Hispanic males. Third, lifespan inequality increased for Hispanic and white people, but decreased slightly for Black people. Fourth, the pandemic shifted racial/ethnic mortality differentials in favor of white people: narrowing the Hispanic advantage and widening the Black disadvantage. Our results provide a comprehensive assessment of mortality trends to inform policies targeting inequalities.

## Main text

The COVID-19 pandemic has taken an unprecedented toll on mortality around the world. Most high-income countries experienced life expectancy losses in 2020 (1–4), and many continued to experience declines in 2021 (5). The United States of America (USA) saw its largest drop in life expectancy (1.7 years for females and 2.1 for males) in recent history (1), with COVID-19 deaths accounting for most of the decline for both females and males (6). Early data showed uneven impacts of the pandemic by race/ethnicity in the USA. The Center for Disease Control (CDC) estimates that between 2019 and 2020, life expectancy decreased by 3 years for the Hispanic population and by 2.9 years for the non-Hispanic Black (henceforth: Black) population, compared to a 1.2-year decline for non-Hispanic white (henceforth: white) people (6). The decrease was largest among Hispanic males (3.7 years) followed by Black males (3.3 years), and smallest among white females (1.1 years). These findings are consistent with early studies projecting a disproportionate impact of the pandemic on life expectancy among race/ethnic minorities (7). It is important to monitor these disparities which reflect underlying inequalities that are often magnified during a public health crisis (8). Both direct deaths from COVID-19 infection (9) as well as indirect deaths from other causes likely disproportionately affected race/ethnic minorities during the pandemic because of the social and economic disadvantages of historically marginalized populations in the USA (6, 10).

This study provides a comprehensive analysis of mortality changes across racial/ethnic groups in the USA before and during the first year of the pandemic. It contributes to the evidence by: analyzing recent trends in life expectancy, average years of life lost, and lifespan inequality from 2010 to 2020 separately for Black, Hispanic, and white populations; 2) identifying the ages and causes of death driving recent changes, including deaths from cardiovascular diseases, respiratory diseases, infectious and parasitic diseases, “deaths of despair” (i.e., suicide-, drug-, and alcohol-related mortality), cancers, accidents, and COVID-19; and 3) comparing race/ethnic gaps in these outcomes before and during the pandemic.

In the years before the pandemic, life expectancy in the USA followed atypical trends of stagnation that have not been observed in most high-income countries (6, 11, 12). These trends have been marked by worsening working-age mortality due to increased drug-related causes of death (13–17), as well as increased deaths from cardiovascular disease at middle and later ages (18). Life expectancy is consistently higher for the white population relative to the Black population, although the gap between Black and white people narrowed from 5.7 years in 2000 to 3.8 years in 2010 (19). This convergence is partly due to relative improvements in mortality from heart diseases, HIV/AIDS, accidents, and cancer (20, 21). By contrast, the Hispanic population had higher life expectancy than the white population throughout the pre-pandemic period, attributable to lower mortality from cancer, cardiovascular diseases, diabetes, chronic respiratory diseases, perinatal conditions, as well as “deaths of despair”. Early evidence suggests that the pandemic has widened the white-Black gap in life expectancy, while reducing the Hispanic advantage (6). However, less is known about which ages and causes of death drove these changes (10).

The COVID-19 pandemic has directly and indirectly affected multiple causes of death. For example, delays in treatment may have increased mortality from cancers (22), or avoidance of hospitals for fear of infection may have increased mortality from acute cardiovascular events (23). COVID-19 is also associated with elevated risk of cardiovascular events and diabetes in the months following infection (24, 25). Crucially, the impact of these changes likely varies across race/ethnic groups, due to differences in socioeconomic resources, rates of health insurance and access to health care (7). Recent findings show that, while COVID-19 death rates were highest in the Hispanic population, Black people experienced exceptionally large increases in mortality from heart disease, diabetes, and external causes of death (10).

So far, research on the differential impact of the pandemic on mortality across race/ethnic groups has mainly relied on estimates of overall life expectancy (6, 7, 26, 27) and standardized death rates (10). While life expectancy is a widely-used and important indicator for studying population health and mortality, it is an average measure which conceals population variability and inequality (28–31). Lifespan inequality is an important measure because it captures a fundamental type of inequality: variation in length of life (32). Two populations that share the same life expectancy could experience differences in the variation around the timing of death. For instance, a high lifespan inequality measure would suggest that deaths occur within a wider age range, while a low measure of lifespan inequality would suggest a narrower age range. Hence, lifespan inequality, measured as the spread of ages at death in a population (e.g. standard deviation), reflects how predictable length of life is at the individual level, and it underlies how uneven mortality improvements are at the population level (32, 33).

Black Americans not only experience shorter life expectancy compared to Hispanic and white people, but also have less predictable lifespans, with higher lifespan inequality (34, 35). The impact of the pandemic on USA lifespan inequality is currently unknown. Evidence from England & Wales shows that both lifespan inequality and life expectancy decreased during 2020 because mortality was concentrated at older ages (36). However, in the USA, life expectancy losses during the pandemic have been driven by increases in mortality both at older and working ages (1, 5, 37). Previous studies show that increased midlife death rates increase lifespan inequality (38–40).

A complementary indicator to life expectancy is ‘average years of life lost’ (AYLL) (41). This refers to the average years of life lost between birth and an upper age-limit, often 95 years, from a synthetic cohort experiencing death rates in a given year throughout their lifespans. For example, if individuals between birth and age 95 live on average 80 years, then there are 15 years of life lost. While other indicators of years of life lost simply add up estimated remaining life expectancies among observed deaths (42, 43), AYLL is comparable across populations and over time, and is not affected by population age structure (44). This indicator enables researchers to quantify the burden of specific causes of death in a given year in a comparable way between populations (45). Using life expectancy, lifespan inequality and AYLL, we comprehensively quantify the unequal impact of age- and cause-specific mortality before and during the first year of the pandemic across race/ethnic groups in the USA.

### Life expectancy, lifespan inequality and years of life lost

Life expectancy for both females and males stagnated in the second decade of the 21st century for all race/ethnic groups, but particularly for white people (Figure 1, panel A). Black and Hispanic females saw small improvements of only six months from 2010-2019, from 77.6y to 78.3y and from 84.6y to 85.3y, respectively. Black males had the lowest life expectancy throughout the period - 71.4y in 2019. Lifespan inequality, measured by the standard deviation of ages at death (see *Methods*), increased in 2010-2019 for all groups and was highest in the Black population. This finding means that the Black population has a double burden compared to white and Hispanic people. Not only are their lives shorter, they are also more unpredictable. Average years of life lost (AYLL) below age 95 varied from 11.7y for Hispanic females to 24.8y for Black males, with little-to no improvements over this period. Graphically, AYLL is the total colored area above the survival (probability of surviving) curve and can be attributed to specific causes of death. Deaths from cardiovascular disease (CVD) accounted for the largest individual share of life lost in 2010 and 2019 (between 25% and 30%) (Figure 1, panel B), followed by cancer (more than 20%). In 2019, “deaths of despair” accounted for 10+% of AYLL in males, with the biggest impact for White males.

**Figure 1.**
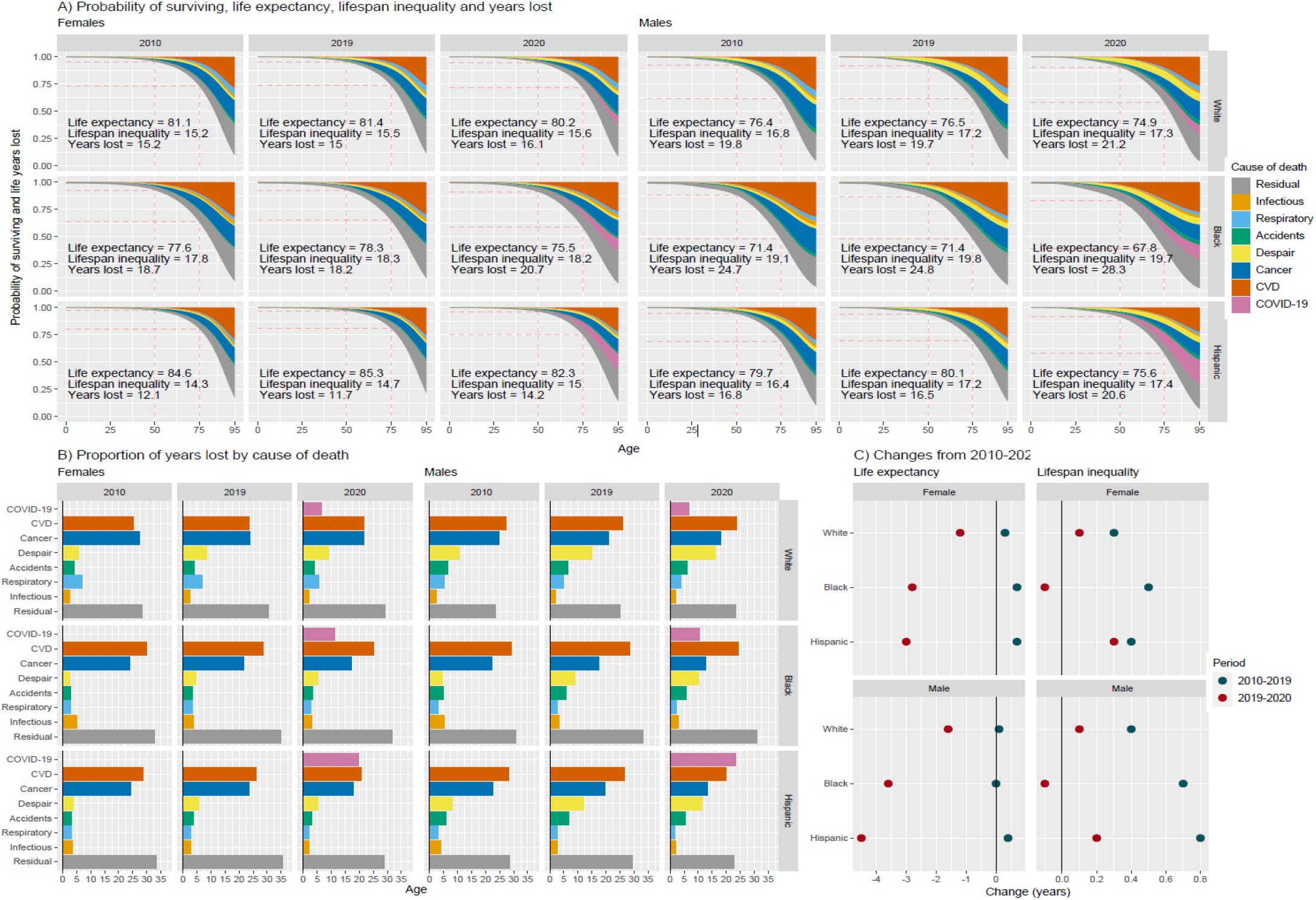
A) Probability of surviving and average life years lost by cause of death (colored areas above), life expectancy, lifespan inequality, and years of life lost by year, sex and ethnic/racial group. B) Proportion of life years lost by cause of death. C) Changes in life expectancy and lifespan inequality.

In 2020, life expectancy decreased for all groups (Figure 1, panel C). Hispanic and Black males saw the largest drops (4.5y and 3.6y respectively), while their female counterparts experienced losses of almost 3y. White females and males lost 1.2 and 1.5y of life expectancy, respectively. Lifespan inequality increased slightly for Hispanic and White populations, but decreased slightly for the Black population. Premature mortality at younger ages is translated into increased lifespan inequality, but increases in mortality at older ages may decrease lifespan inequality. Hence, the divergent patterns in lifespan inequality across groups suggest varied impacts of the pandemic by age. AYLL also increased substantially for all groups. The largest increase in AYLL in 2020 was found in Hispanic males (4.1y), where COVID-19 contributed the most among specific causes (almost 25% of life lost) (Figure 1, panel B), followed by Black males (3.5y) where COVID-19 accounted for more than 10% of the total years lost.

### Trends in life expectancy and lifespan inequality by age and causes of death

The stagnation of life expectancy in the U.S. from 2010-2019 reflected changing age-patterns of mortality in certain causes of death. Improvements in cancer mortality, predominantly in ages 50-90, contributed to increases in life expectancy across all groups in this period (Figure 2). The biggest gains from cancer improvements were observed for Black females and males (0.42y and 0.69y), and white males (0.48y) (see supplementary figure S2). Improvements in cardiovascular mortality also contributed positively to life expectancy in 2010-2019, especially for females (0.24y for white, 0.40y for Black, and 0.47y for Hispanic females). However, this progress was reversed by increased mortality from “deaths of despair”, with males in all race/ethnic groups losing a staggering half a year or more from “deaths of despair” from 2010-2019. Losses from “deaths of despair” were concentrated in younger ages, with 30-39 year-olds suffering the greatest life expectancy losses in all race/ethnic groups. While females also lost life expectancy from “deaths of despair” in 2010-2019, the magnitude was smaller compared to males (0.16y).

**Figure 2.**
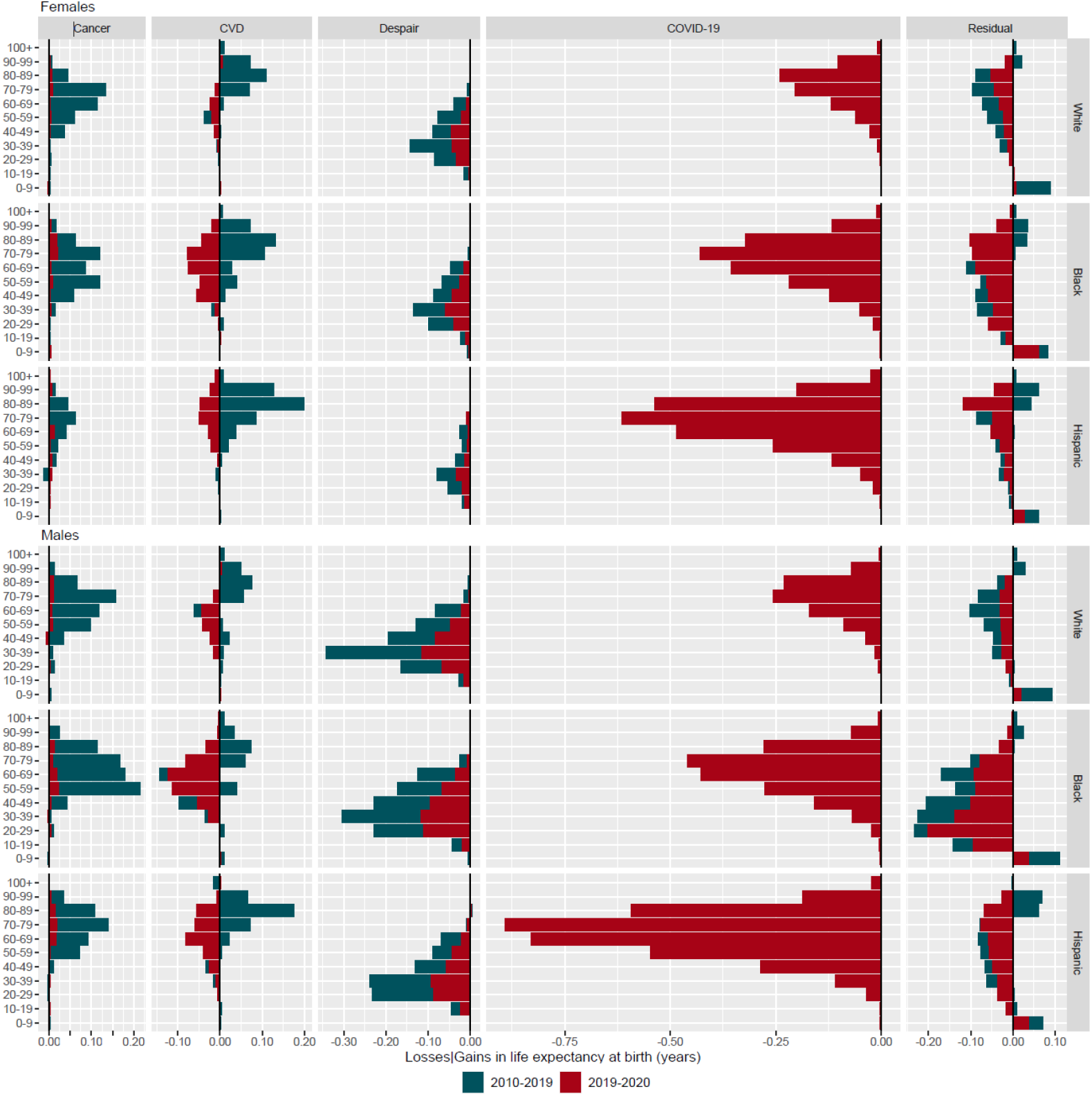
Contributions by age and causes of death to changes in life expectancy in 2010-2019 and 2019-2020 by race/ethnic groups and sex. Note: see supplementary figure S1 for infectious and respiratory diseases.

The pandemic induced changes in the patterns of mortality by cause of death (Figure 2, red color). In 2020, small cancer improvements translated into negligible gains in life expectancy, while deaths from cardiovascular mortality, “deaths of despair”, and residual mortality increased dramatically. These changes translated into significant overall losses of life expectancy. For example, “deaths of despair” contributed to life expectancy declines of 0.35y for white, 0.46y for Black, and 0.33y for Hispanic males from 2019 to 2020, on top of already persistently high levels of “deaths of despair” in the previous decade. As expected, the largest contributions to life expectancy losses were from deaths due to COVID-19. Official COVID-19 deaths translated into losses of 0.88y for white, 1.78y for Black, and 3.50y for Hispanic males. The corresponding losses among females were 0.78y, 1.65y and 2.31y. In contrast to other high income countries, COVID-19 deaths at working ages contributed significantly to life expectancy declines. For example, among Hispanic and Black males, mortality below age 60 accounted for 29.9% and 31.9% of the total decline due to COVID-19 in life expectancy, compared to 18.4% for white. For females, the corresponding proportions were 14.7%, 27.2% and 21.0% for white, Black and Hispanic people, respectively.

Increased mortality at any age translates into life-expectancy losses. However, for lifespan inequality to increase, mortality deterioration needs to be concentrated in younger ages. This means that worsening mortality at older ages, above a threshold age usually very close to life expectancy, may lead to decreased lifespan inequality (46–48). For example, lifespan inequality decreased in England & Wales in 2020 due to worsening death rates at older ages during the pandemic. Causes of death that explain increases or decreases in life expectancy, are often not the same driving decreases or increases in lifespan inequality (49, 50). In the USA lifespan inequality increased for all groups between 2010 and 2019. Increases were largely driven by mortality from “deaths of despair” below age 50 and improvements in cardiovascular disease mortality at ages 70+ (see supplementary figure S3). In 2020, lifespan inequality continued to increase for white and Hispanic people, while the Black population experienced small declines. Increased mortality at older ages from COVID-19 contributed to decreasing lifespan inequality among Black and white people, but not enough to see overall declines in lifespan inequality among Blacks. Among Hispanic males COVID-19 deaths contributed to increasing lifespan inequality because they were concentrated at younger ages, increasing uncertainty in lifespan. “Deaths of despair” continued to increase lifespan inequality in 2020 for all groups.

### Life expectancy and lifespan inequality gaps between race/ethnic groups

Hispanic people have consistently had the highest life expectancy among all race/ethnic groups in the USA, and Black people the lowest. Between 2015 and 2019, life expectancy gaps between the Hispanic and white population remained stable at around 3.8y for both females and males (figure 3). The gap between white and Black males increased from 4.6y to 5y in 2015-2019, while for females it increased minimally from 2.9y to 3.1y. Figure 3 shows age- and cause-specific contributions to the gap in Hispanic-white and white-Black life expectancy in 2015, 2019 and 2020 (for Hispanic-Black comparisons see appendix figure S4). In 2015 and 2019, the Hispanic-white gap was largely explained by a Hispanic advantage in mortality at adult ages (20 and older) for both females and males. In these years, the causes of death that contributed the most to the gap were cancer, “deaths of despair”, cardiovascular diseases and residual mortality (see appendix figure S5). In contrast, higher mortality under age ten among Black people contributed substantially to the white-Black gap in life expectancy. Deaths from cardiovascular diseases and residual causes contributed the most to the white-Black gap. The only causes of death less severe in Black compared to white people were “deaths of despair” (negative contributions in Figure 3). This means that if Black males were to experience the same mortality rates of “deaths of despair” as white males, the gap would be even larger.

**Figure 3.**
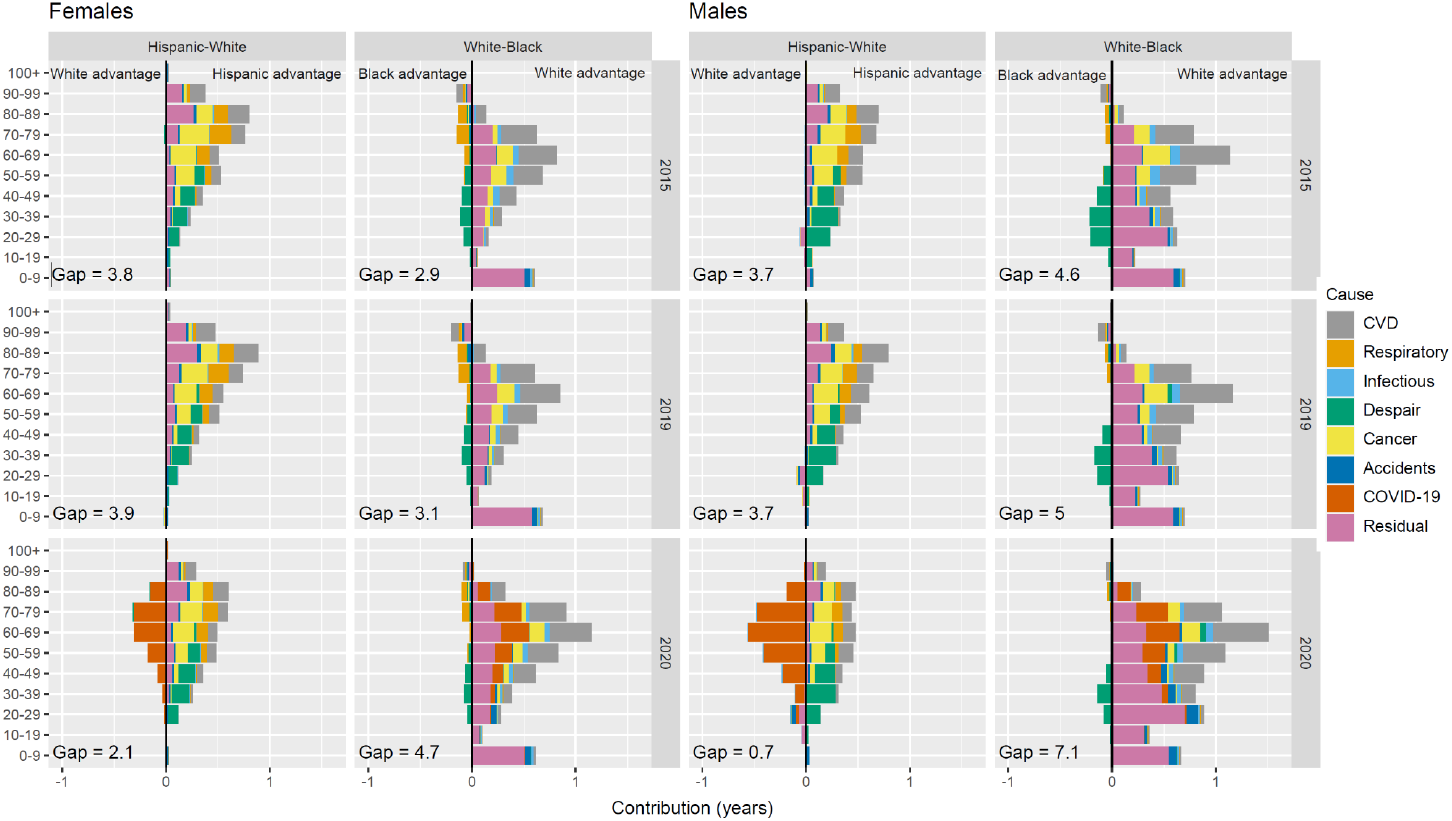
Contributions by age and causes of death to race/ethnic gaps in life expectancy in 2015, 2019 and 2020 by sex.

In 2020, over the course of the pandemic, the Hispanic-white life expectancy gap narrowed to 2.1y for females and 0.7y for males, a decline of 1.7y and 3.1y, respectively. Conversely, the white-Black gap widened to 4.7y for females and 7.1y for males, an increase of 1.6y and 2.1y, respectively. The decrease in the Hispanic-white gap was driven by the disproportionate impact of COVID-19 deaths at working and older ages among Hispanic people, and doesn’t reflect improvements in white mortality (negative values in Figure 3). The white disadvantage accounting for the gap was evident at ages below 60 from “deaths of despair”, and from respiratory and cancers at older ages. The increasing white-Black gap in 2020 was largely explained by increased COVID-19 mortality above age 40, together with cardiovascular mortality. Other non-COVID cause-specific contributions to the white-Black gap remained similar to previous years. These trends in mortality also affected lifespan inequality (see appendix figure S6), but to a lesser extent. In 2020, the Hispanic population had the lowest lifespan inequality and the Black population had the highest.

### Outlook and implications

Previous pandemics, such as the 1918 Spanish influenza and the 2009 H1N1 influenza, affected population health and mortality differently across sub-populations (51). Overwhelming evidence, including our own, shows that so far the COVID-19 pandemic has disproportionately affected racial/ethnic minorities in the USA (6, 7, 10, 52). We have provided a comprehensive analysis of mortality profiles for the white, Black and Hispanic populations by age and cause of death in 2010-2019 and 2020, using a set of complementary demographic indicators of average mortality and its variability. Our findings show that the pandemic substantially affected existing mortality differentials in the United States in favor of the white population, narrowing the Hispanic advantage and widening the Black disadvantage in life expectancy and related measures.

One year of pandemic-associated mortality nearly eliminated the Hispanic life expectancy advantage that was consistent over the 2010-2019 period. The declining Hispanic mortality advantage was predominantly driven by a greater concentration of COVID-19 deaths among working-age Hispanic people compared to the white population, a finding that has also been documented elsewhere (6, 10). This is bolstered by our finding that lifespan inequality increased for the Hispanic population, indicating greater variability in ages at death among Hispanic people in the USA. These patterns can be attributed to higher likelihood of viral exposure (due to employment and housing) and lower access to health care (52). The high burden of COVID-19 mortality in the Hispanic population, especially at working ages, suggests that exposure-related social factors outweighed any protection from previously accumulated health advantages. A key question is whether the Hispanic population will recover its life expectancy advantage in the short, medium or long term despite its socioeconomic disadvantage relative to the white population. This phenomenon is known in demographic research as the “Hispanic paradox” (64). Given that the reduction in the Hispanic-white life expectancy gap was largely explained by direct deaths due to COVID-19, vaccination rollout and age-specific uptake may be most relevant in the short term. Recent vaccination data suggests that Hispanic people are slightly more likely than white people to have received at least one dose (64% vs. 62%) (53).

In 2020, the persistent white-Black gap in life expectancy widened. This is the result of larger increases in COVID-19 and CVD mortality among Black people at older ages compared to their white peers, a finding that is consistent with previous studies (10). Although some of the CVD deaths in this population may be attributable to more underreporting of COVID deaths for Black people, the increase is dramatic and warrants further investigation. Black people in the USA may have experienced a “syndemic” of COVID-19 and other chronic diseases (51). Our results suggest that, distinct from the Hispanic population, Black people may have been more exposed not only to COVID-19, but also to the indirect consequences of the pandemic, including delayed or missed healthcare (10, 27). In contrast to the Hispanic-white gap, the white-Black gap in life expectancy may remain wider than it was in 2019 in the short-to-medium term, due in part to persistently lower vaccination rates among Black people relative to both White and Hispanic people (57% with at least one dose according to the KFF report). It is plausible that this gap may continue to widen in 2022 because of the proportional increase in the hospitalization of unvaccinated Black adults during the Omicron-period (since December 2021) compared to other race/ethnic populations in the USA (54). Any indirect impacts of the pandemic that led to increases in CVD mortality may also persist longer than acute COVID risks.

Somewhat paradoxically, lifespan inequality decreased slightly for Black people, because the increase in mortality was primarily at older ages. A similar result was documented in England & Wales for 2020 (36). Typically, life expectancy increases are accompanied by decreases in lifespan inequality (28, 30, 31). This means that countries that have achieved higher levels of life expectancy benefit from lower lifespan inequality. However, recent evidence has challenged the uniformity of this association. For example, in Eastern and Central Europe, life expectancy and lifespan inequality moved independent from one another before the dissolution of the Soviet Union due to worsening mortality at younger ages related to alcohol consumption and improvements in mortality at older ages (55). Similarly, groups with lower socioeconomic status in Finland and Spain experienced life expectancy increases and widening lifespan inequalities simultaneously usually due to high midlife mortality (32, 56). Our results are further evidence of the potential decoupling of life expectancy and lifespan inequality in the context of the pandemic, pointing to the importance of the age-profile of mortality during a crisis for changes to the predictability of lifespan.

The impact of the pandemic may be particularly unequal in countries characterized by high social inequality (e.g., as measured by the standard Gini coefficient), such as the USA (51). Evidence from England - a comparatively unequal high-income country - suggests that South Asian and Black people had considerably higher risk of COVID-19 infection, hospitalization, and death than the white population (57, 58). More research aimed at understanding the link between social factors and racial/ethnic differences in the impact of the pandemic is needed. It is also important to expand this research to low- and middle-income countries, where the impact of the pandemic may have been particularly unequal due to a combination of high inequality and less developed healthcare systems. In the USA, prospects for 2021 are pessimistic and inequalities may have worsened. Recent evidence shows that in 2021 life expectancy continued to decrease in the USA, with an estimated further drop of 0.3 months on top of 2020 losses (5). Relative to 2020, 2021 was also characterized by a shift to younger ages at death for COVID-19.

There are some limitations to this study, related to the availability and quality of the data that were analyzed. Many deaths for which COVID-19 was the underlying cause may have been attributed to CVDs, and this misreporting may have disproportionately affected the Black population, driving the increase in CVD deaths in this group (10). Finally, because of data limitations, this study examines three race/ethnic groups, and does not consider other minorities including American Indians and Alaska natives, Asians, and other race/ethnic groups.

Despite these limitations, this study offers several important lessons for scientists and policymakers. The pandemic has affected mortality unequally. COVID-19 mortality hit Black and Hispanic Americans especially hard, exacerbating existing white-Black disparities and nearly eliminating the Hispanic mortality advantage. Our findings of higher Hispanic mortality at working ages are consistent with previous work suggesting increased risk of COVID-19 exposure for this group due to working conditions, housing, and access to preventive information (27, 52). The widening of the white-Black gap in life expectancy is particularly concerning because, as estimated by Wrigley-Field (8), the difference between Black and white mortality pre-pandemic was already larger than the overall impact of the COVID-19 pandemic on white mortality.

Overall, our findings confirm the wide-ranging impact of the COVID-19 crisis on racial/ethnic differences in mortality in the USA. While the SARS-CoV-2 virus itself does not discriminate, the social environment shapes risk of infection and death in ways that reflect historical inequalities. Future work should continue to examine the direct impact of COVID-19 but also the impact of pandemic social and economic disruptions on racial/ethnic differences in health. Besides risk of increased socioeconomic disadvantage during the pandemic, Hispanic and Black people also suffered much higher levels of bereavement and loss from COVID-19 in their own families(59), contributing to significant trauma and stress that may have long term health effects. Our study provides the first comprehensive assessment of the impact of the COVID-19 pandemic on racial/ethnic gaps in mortality, to ultimately inform research and policy interventions targeting such inequalities.

## Materials and Methods

### Data

We use data from the publicly-available United States multiple cause of death files, from the National Vital Statistics System division of the National Center for Health Statistics (https://www.cdc.gov/nchs/data_access/vitalstatsonline.htm#Mortality_Multiple), and yearly population estimates compiled by the Surveillance, Epidemiology, and End Results Program (https://seer.cancer.gov/popdata/download.html). Data are restricted to USA residents in the years 2010-2020. Non-Hispanic white (white), non-Hispanic Black (Black), and Hispanic/Latino (Hispanic) populations are analyzed, and all other race/ethnic groups were excluded because of small data counts. We code six causes of death: cardiovascular diseases (CVD), respiratory diseases, infectious and parasitic diseases (IPD), “deaths of despair” (suicides, drug- and alcohol-related deaths), cancers, accidents (excluding those from “deaths of despair”), and COVID-19. While “deaths of despair” encompass three causes of death, driven by underlying phenomena that are likely independent (13, 14), evidence suggests that all three causes were rising before and during the pandemic. As such, we combine them here. Specific International Classification of Disease (ICD)-10 codes are listed in Table 1.

**Table 1.** ICD codes for groups of causes of death

| Category | ICD-10 Codes |
| --- | --- |
| CVD | I00-I78 |
| Respiratory Illness | J40-J46 |
| Infectious and Parasitic Diseases | A00-A09, A16-A44, A48-A99, B00-B09, B15-B99, D86.9, G02, G14, H32, I32, I39 J17, K90.8, L44.4, L94.6, M02.3, M35.2, M66, N34.1, R11.1 |
| Despair | F10-F16, F19, K70, K73-K74, U03, X40-X45, X64-X85, Y10-Y15, Y87 |
| Cancer | C00-C99 |
| Accidents | V00-V99, W00-W99, X00-X59, Y85-Y86 (excluding, despair causes) |
| COVID | U07.1, U07.2 |
| Residual | All else |

### Life table construction

Life tables for all race/ethnic groups by sex for the period 2010-2020 were constructed following standard demographic techniques consistent with a piece-wise constant hazard model using all-cause mortality by single ages with the last age interval grouping deaths at ages 100+ (60).

### Life expectancy, average years of life lost and lifespan inequality

From the life tables, we retrieved life expectancy at birth, life years lost, and calculated lifespan inequality as an indicator of how spread out are ages at death.

Life expectancy at birth is a summary indicator of population health and mortality. It expresses the average number of years a synthetic cohort of newborns is expected to live if they were to experience the mortality rates observed in a given year (61). It is not affected by population size or age-structure, which makes it a preferred indicator for comparisons over time and between populations.

Average life years lost (AYLL) is a complementary indicator to life expectancy. It refers to the average years lost of a synthetic cohort due to mortality in specific age-groups or causes of death (41). For example, if individuals live on average 80 years between birth and age 95, then AYLL equals 10 years. In this analysis we set the upper limit of 95 to quantify AYLL. This strategy has been used previously to characterize mortality in contexts where subpopulations experience different mortality profiles, e.g., population with and without mental health disorders (44).

Lifespan inequality refers to variation in ages at death and is a marker of heterogeneity in mortality at the population level and reflects uncertainty in lifetimes at the individual level (32). The greater lifespan inequality, the more unpredictable lifespans are in a population. We measure lifespan inequality with the standard deviation of the age-at-death distributions. Several indicators exist to measure lifespan inequality including the life table entropy, the Gini coefficient, or life disparity, and all are highly correlated when measured from birth (62), which suggest that our results would not change on the basis of the indicator chosen.

### Decomposition of life expectancy by age and cause of death

In order to disentangle age- and cause-specific effects over time and between groups for life expectancy and lifespan inequality we used the linear integral decomposition method (63), a state-of-the-art method that allows us to decompose the difference of two values of life expectancy or lifespan inequality by age and cause of death, which has been implemented previously for this type of analysis (e.g. ref. (1)).

## Data Availability

The replication files for this paper include customized functionality written in the R statistical programming language. The code, and all harmonized input and output data pertaining to our analysis, is hosted both on Zenodo (a general-purpose open-access repository developed under the European OpenAIRE program and operated by CERN) at https://zenodo.org/record/6402403, and on GitHub https://github.com/jmaburto/ex_USA_racial-ethnic_differences.

## Authors contributions

Conceptualization: JMA, AMT, GF, JBD

Data curation: AMT

Formal analysis: JMA

Methodology: JMA

Software: JMA, AMT

Visualization: JMA

Writing – original draft: JMA

Writing – review & editing: JMA, AMT, GF, JBD

## Data and materials availability

**Supplementary information is available for this paper**: Figs. S1 to S6.

## Supplementary Information

**Figure S1.**
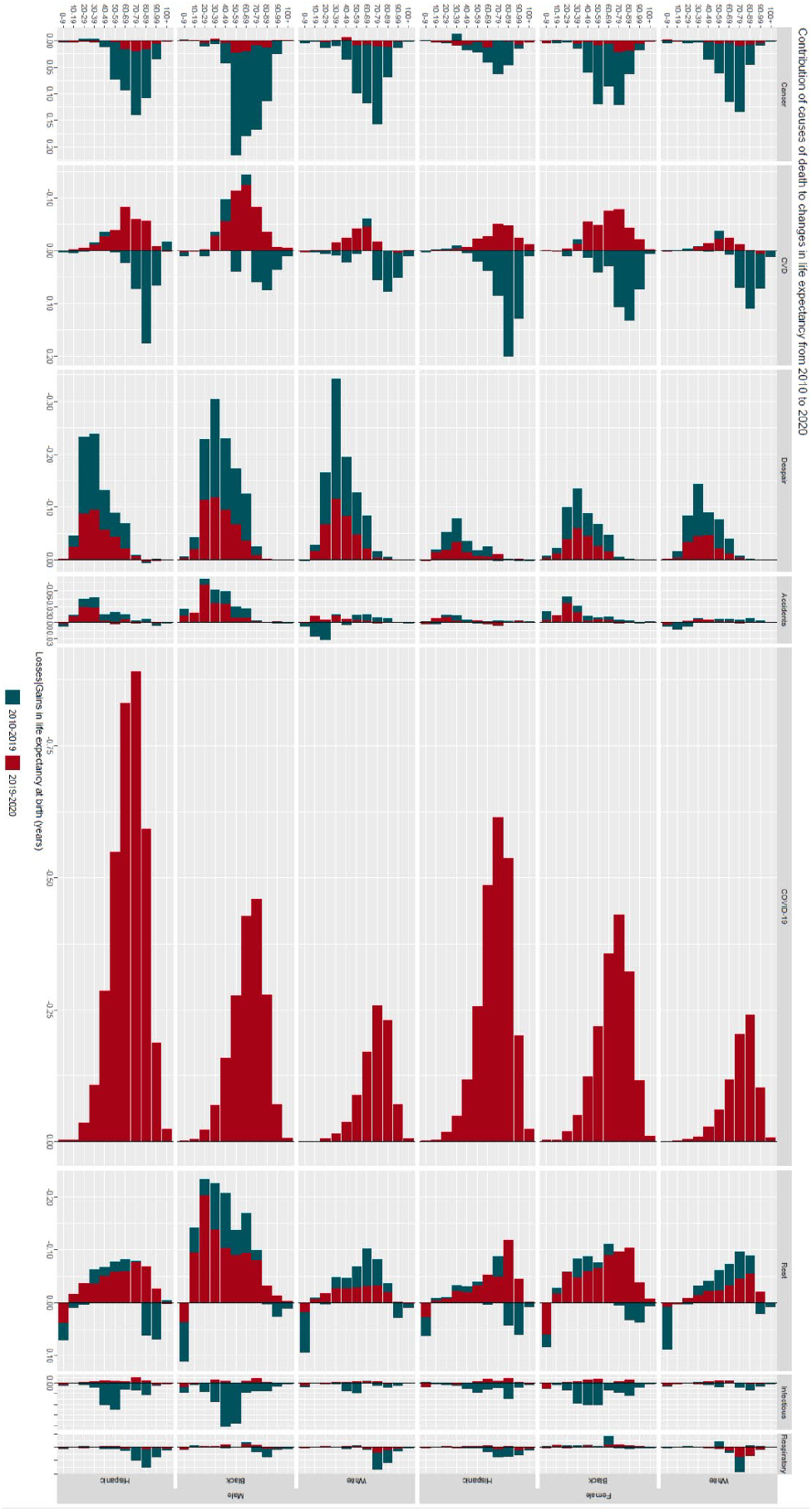
Contributions by age groups and causes of death to changes in life expectancy in 2010-2019 and 2019-2020 by racial/ethnic groups and sex.

**Figure S2.**
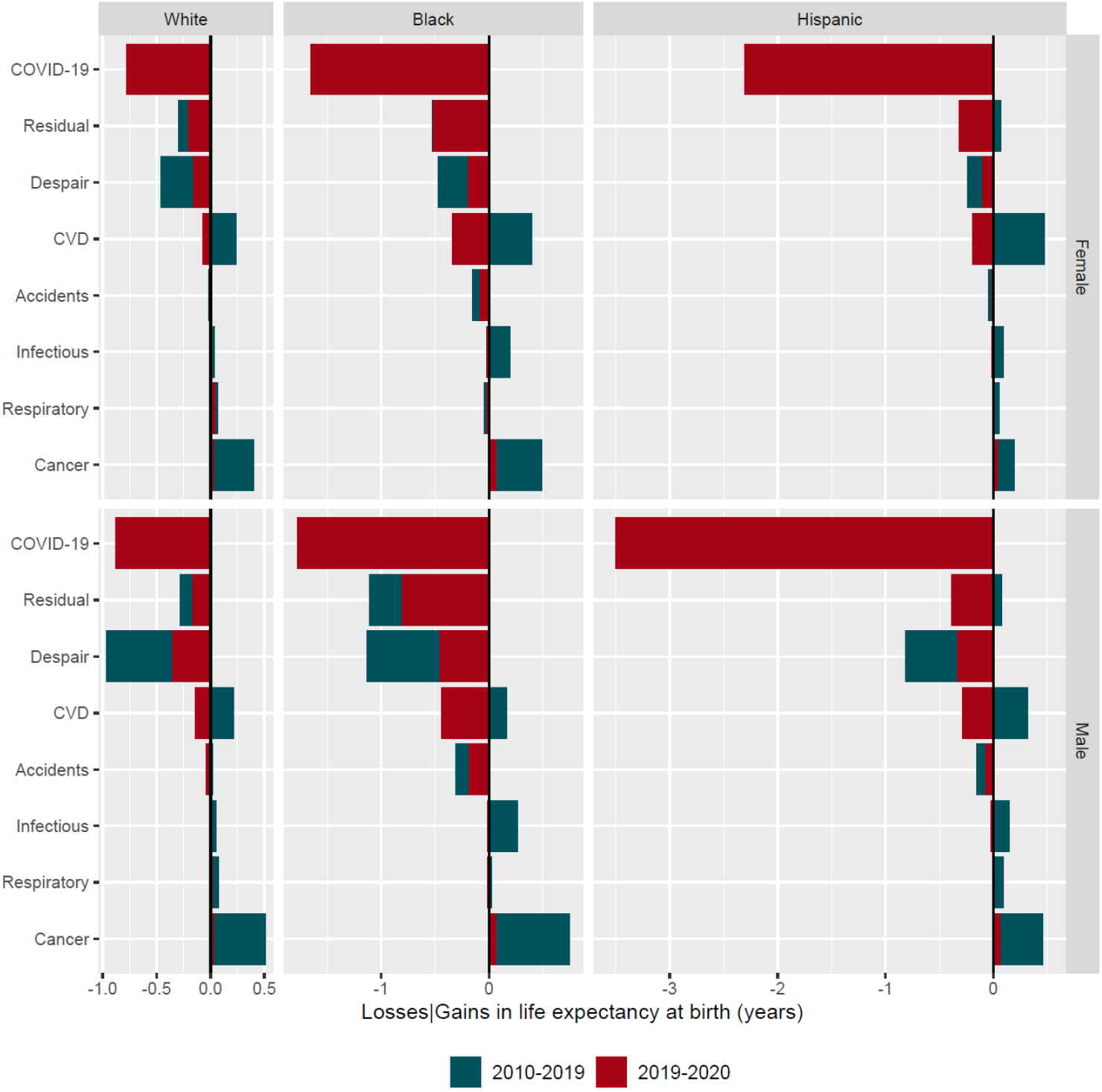
Contributions by causes of death to changes in life expectancy in 2010-2019 and 2019-2020 by racial/ethnic groups and sex.

**Figure S3.**
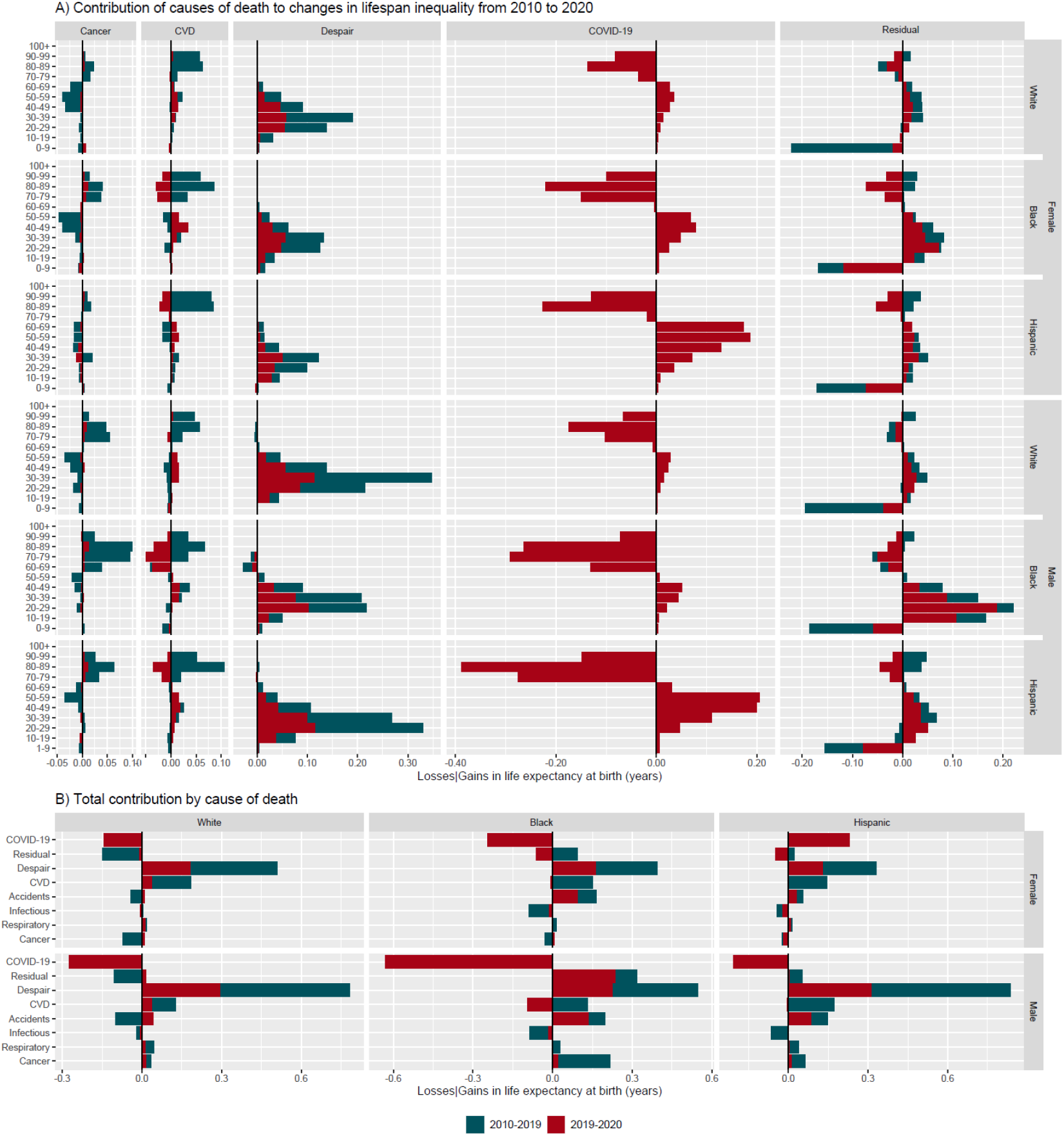
Contributions by causes of death to changes in lifespan inequality in 2010-2019 and 2019-2020 by racial/ethnic groups and sex.

**Figure S4.**
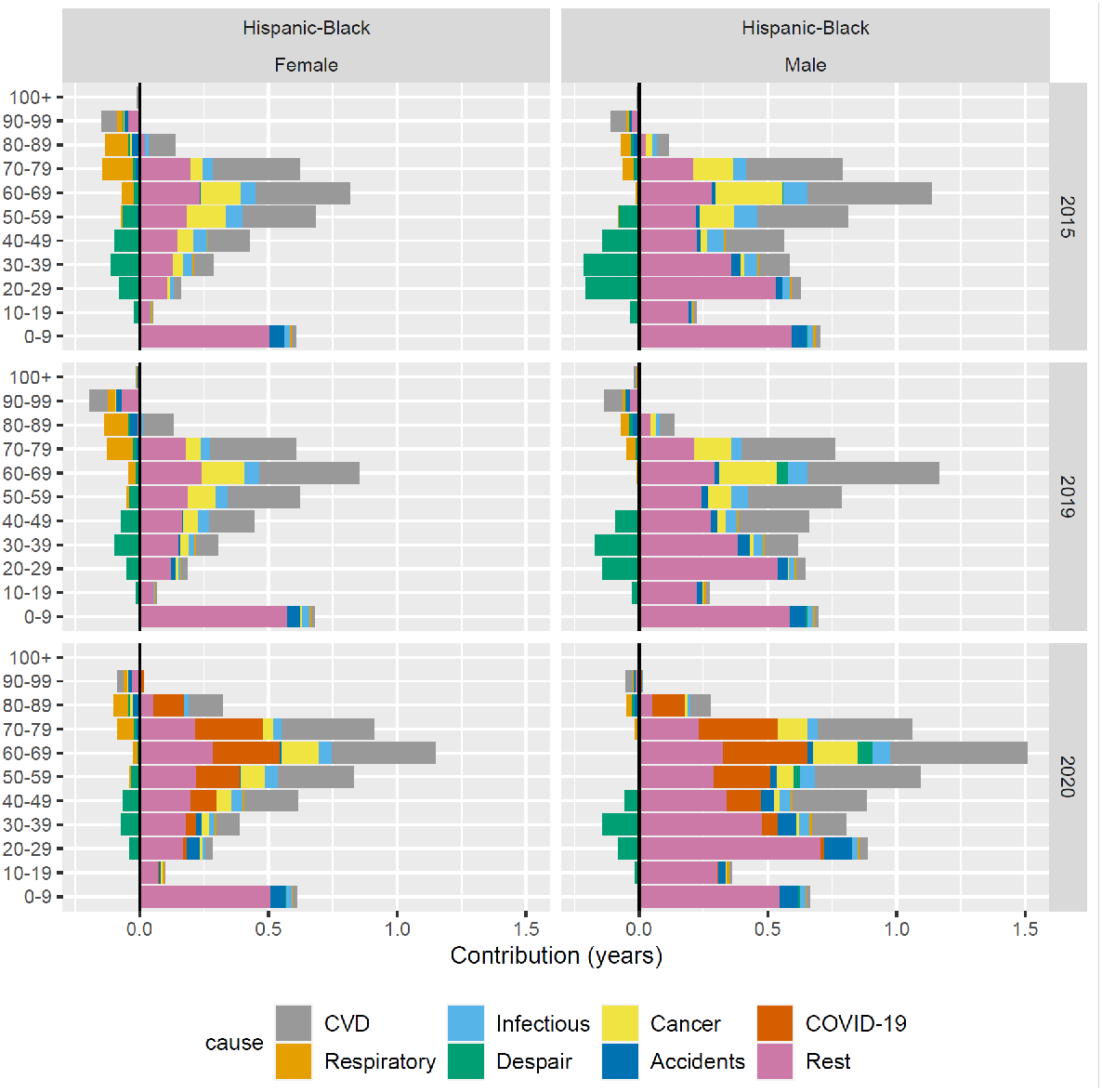
Results as shown in Figure 3 for Hispanic-Black difference in life expectancy.

**Figure S5.**
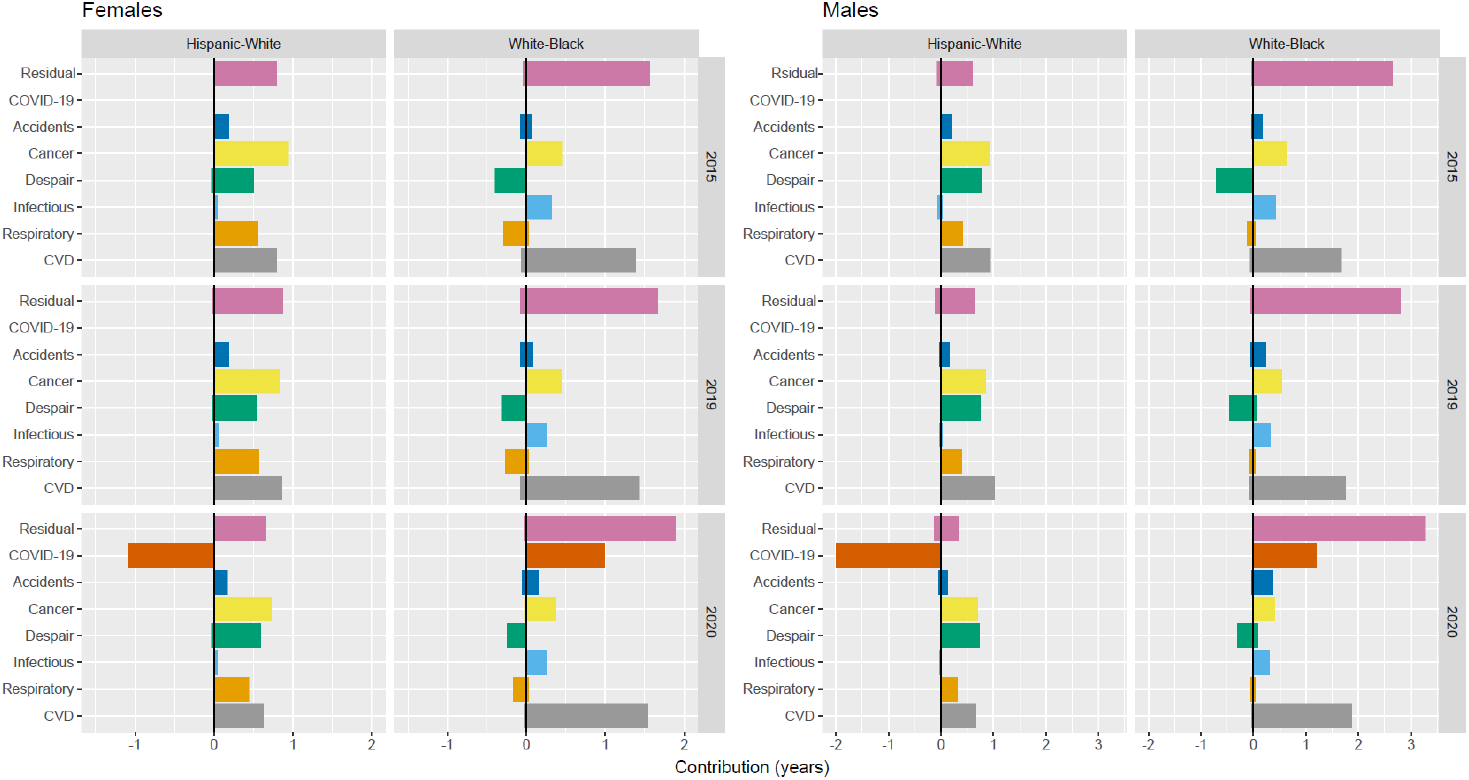
Contributions by causes of death to ethnic/racial gaps in life expectancy in 2015, 2019 and 2020 by sex.

**Figure S6.**
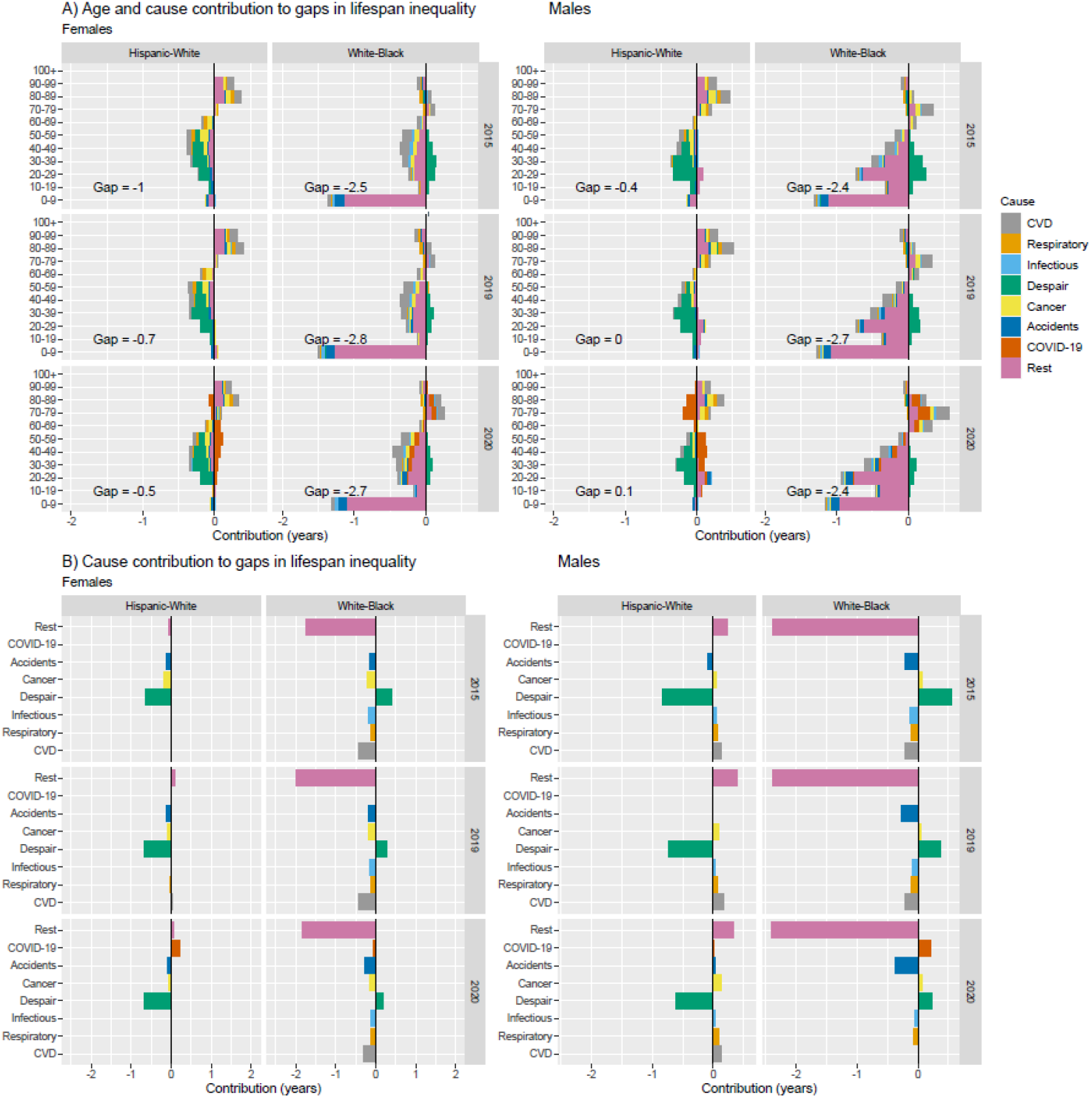
Contributions by age and causes of death to ethnic/racial gaps in lifespan inequality in 2015, 2019 and 2020 by sex.

